# Implementing Learning Health System Principles to Advance the Evaluation and Treatment of Clinical High-Risk for Psychosis

**DOI:** 10.1101/2025.05.09.25327345

**Authors:** Cheryl Y.S. Foo, Lauren Utter, Abigail L. Donovan, Corinne Cather, Daphne J. Holt, Jacqueline A. Clauss

**Affiliations:** Psychosis and Clinical Research Program, Department of Psychiatry, Massachusetts General Hospital, Boston, MA; Department of Psychiatry, Harvard Medical School, Boston, MA; Center of Excellence for Psychosocial and Systemic Research, Department of Psychiatry, Massachusetts General Hospital, Boston, MA; Maryland Psychiatric Research Center, Department of Psychiatry, University of Maryland Medical School, Catonsville, MD

**Keywords:** clinical high-risk for psychosis, learning health system, transdiagnostic factors, identification, treatment

## Abstract

**Background:** Early intervention for individuals with clinical high-risk for psychosis (CHR-P) aims to prevent the onset of serious mental illnesses like schizophrenia, but models for specialized care remain underdeveloped. We describe the development, feasibility, and value of a CHR-P evaluation and treatment program built on learning health system principles. We demonstrate that referral and intake clinical assessment data can improve the characterization and identification of individuals with CHR-P.

**Methods:** The Resilience Evaluation-Social Emotional Training (RE-SET) Program, developed as part of a continuum of psychosis care programs at Massachusetts General Hospital, systematically collects clinical data at referral, intake evaluation, and during treatment. In this study, at referral, providers, caregivers, and/or patients reported on the patient’s psychiatric history. Patients for whom there were concerns for attenuated psychosis were eligible for evaluation. Prior to diagnostic evaluation, patients completed self-report measures of a broad range of psychiatric symptoms. Patients were evaluated with the Structured Interview for Psychosis-Risk Syndromes (SIPS) to determine if they met CHR-P criteria. Using referral and intake data from 118 help-seeking individuals, we performed univariate chi-square or independent samples t-test to identify factors associated with CHR-P syndrome.

**Results:** Ninety-nine (99) individuals with complete referral data were included in the analysis (mean age: 17.8 years; SD: 4). Almost a quarter (24.2%) met CHR-P criteria on the SIPS. Referred individuals presented with high rates of psychiatric comorbidity, previous psychiatric treatment, and functional impairment across multiple domains. Significant predictors (p<.05) of CHR-P syndrome included: history of autism spectrum disorder, endorsing more than one psychotic symptom, reduced sleep duration, and more severe cognitive and behavioral difficulties.

**Conclusions:** Identification of CHR-P may be improved with greater attention to developmental history, psychotic symptoms, sleep disturbance, and cognitive and behavioral difficulties. Integrating routine assessment into clinical care provides data-driven opportunities to improve the identification and treatment of CHR-P.

## Introduction

The clinical high-risk for psychosis (CHR-P) syndrome was identified to promote early detection and intervention among individuals at elevated risk for developing a psychotic disorder and other serious mental illnesses, potentially improving outcomes and preventing the onset of these disorders (Catalan, Salazar de Pablo, et al., 2021). CHR-P individuals are characterized by the presence of subthreshold or attenuated psychotic symptoms, brief intermittent psychotic symptoms, or a family history of psychosis, diagnosed by a structured clinical interview (Fusar-Poli et al., 2016; Woods et al., 2019, 2023). Besides having up to 20-fold increased risk for psychotic disorders (Fusar-Poli et al., 2012), CHR-P individuals are also at risk for poor long-term mental health and functional outcomes, including severe mood disorders, anxiety disorders, lower educational attainment, and lower functioning (Addington et al., 2011; Salazar De Pablo et al., 2022). As such, specialized services for individuals with CHR-P have emerged globally as innovative models for early detection, comprehensive assessment, and targeted intervention to modify illness trajectories of a broad spectrum of emerging severe psychopathology, including both psychotic and non-psychotic disorders (McGorry et al., 2018; Patel et al., 2007; Phillips et al., 2002).

Despite these advances, significant gaps persist in our understanding of how to accurately identify and tailor treatment for a heterogenous CHR-P population. Approximately 80% of individuals who present for CHR-P evaluation have at least one comorbid diagnosis, most commonly mood and anxiety disorders (Addington et al., 2017; Solmi et al., 2023; Woods et al., 2018). Attenuated psychotic symptoms, the hallmark symptoms of CHR-P, are observed in other mental health conditions, and can be difficult to distinguish from symptoms of autism, obsessive-compulsive disorder, and severe anxiety disorders (Addington et al., 2011; Simon et al., 2014; Solomon et al., 2011; Welikson & Guvenek-Cokol, 2023). CHR-P individuals also have different illness and recovery trajectories (Addington et al., 2011); approximately 20% of individuals with CHR-P progress to a psychotic disorder within two years of CHR-P diagnosis, and an additional 10% of individuals develop a psychotic disorder over four or more years (Salazar De Pablo et al., 2021). The substantial proportion of CHR-P individuals who do not progress to a psychotic disorder continue to have other serious psychiatric morbidity (Addington et al., 2011; Salazar De Pablo et al., 2022). Given the heterogenous clinical presentations and trajectories of CHR-P individuals, there are no consensus pharmacotherapy guidelines for individuals with CHR-P (Catalan, Salazar de Pablo, et al., 2021). While there are standalone psychosocial treatments (e.g., cognitive behavioral therapy, family focused therapy) that are effective in reducing symptom burden and functional impairment in CHR-P individuals (Thompson et al., 2015), no evidence-based treatment model exists for CHR-P that is akin to the gold-standard coordinated specialty care for early psychosis treatment (Dixon et al., 2015; Fusar-Poli et al., 2019; Robinson et al., 2022). These challenges highlight the critical need for research and service innovation that can inform and improve CHR-P identification and treatment.

A learning health system framework offers a compelling paradigm for the development and implementation of specialized CHR-P programs that can address these challenges (*Learning Health System Series - NAM*, 2024; McGinnis et al., 2021). A learning health system seamlessly integrates research and clinical practice, where continuous knowledge generation and application to quality improvement occurs in tandem with care delivery. This is often achieved by a clinical infrastructure that incorporates systematic and standardized measurement, which can simultaneously serve clinical decision-making and research purposes. Clinical outcomes are systematically monitored, and services are routinely evaluated and refined. CHR-P programs built on these learning health system principles have the potential to offer insights into improving accurate identification and assessment of CHR-P syndrome as well as optimized intervention strategies based on illness profiles and trajectories.

In this paper, we describe the development, feasibility, and value of a specialized CHR-P evaluation and treatment program located in an academic hospital in Massachusetts, USA, built upon learning health system principles. We illustrate how systematic clinical assessment data collection can improve identification and characterization of CHR-P syndrome. Improving identification of CHR-P individuals can facilitate more timely referrals and reduce evaluation burden, as well as provide indications for treatment approaches. Specifically, with these data, we aimed to identify: 1) the demographic and clinical profile of young persons referred to a CHR-P program; and 2) clinical characteristics at referral and intake that were associated with CHR-P syndrome.

## Methods

### Setting

The Massachusetts General Hospital (MGH) Resilience Evaluation-Social Emotional Training (RE-SET) Program (https://www.resilienceandprevention.com/re-set) provides diagnostic evaluation and ongoing care for youth aged 12-30 years who report or are suspected to be experiencing attenuated psychotic symptoms, but who do not have an established primary psychotic disorder diagnosis. The RE-SET Program was formed through a partnership between the MGH Resilience and Prevention Program and the MGH Psychosis Clinical and Research Program. The Resilience and Prevention Program was founded in 2012 by Dr. Daphne Holt as a research program within the MGH Department of Psychiatry. It was developed to develop, test, and disseminate evidence-based, neuroscience-informed, behavioral interventions for the prevention of psychiatric illnesses in transdiagnostically at-risk youth or adult populations. The Psychosis Clinical and Research Program was established to provide stage-based care across the continuum of psychotic illness, including: RE-SET for CHR-P; a first-episode psychosis program, which provides up to three years of coordinated specialty care for individuals within the past three years of psychotic illness onset; a chronic psychosis program; and a psychosis consultation service. The PCRP outpatient programs are co-located in one office suite at MGH, allowing for shared resources and research activities. Staff in RE-SET provide care in the other programs, allowing for coordination across programs and the facilitation of timely transitions to the appropriate program based on the patient’s illness stage. By partnering with a psychosocial research program, the RE-SET Program is able to integrate research within clinical care and to test novel interventions and treatments.

Informed by the gold-standard, coordinated specialty care service for first-episode psychosis in the USA (Dixon et al., 2015; Robinson et al., 2022), RE-SET provides team-based care that includes medication management, individual therapy (cognitive-behavioral therapy approaches), family education and support, educational support and advocacy, neuropsychological assessment, and care coordination. RE-SET clinicians are child, adolescent, and adult-trained psychiatrists and psychologists, as well as supervised doctoral-level psychiatry fellows and psychology trainees. Patients can receive up to three years of ongoing care in RE-SET. Clinical care provided by RE-SET is billed to insurance; however, non-billable services have been supported by grants from the Substance Abuse and Mental Health Services Administration (SAMHSA) block grant funding to the Massachusetts State Department of Mental Health. RE-SET is also part of the Massachusetts Psychosis Prevention Project (M3P), a consortium of CHR-P programs in the state that received this block grant funding.

### Ethics

The Mass General Brigham Institutional Review Board approved this study, and deemed the study exempt from the requirement to obtain informed consent, since the data were collected for clinical and quality improvement purposes.

### Evaluation and Measurement-Based Care Procedures

We administered a comprehensive battery of validated self-report and caregiver-report measures to facilitate evaluation of CHR-P and other psychiatric conditions. Diagnostic evaluation consisted of three stages: 1) referral screening, 2) diagnostic evaluation with clinical assessment battery, and 3) Structured Interview for Psychosis-Risk Syndromes (SIPS; CHR-P assessment; Woods et al., 2019). For purposes of this study, “CHR-P individuals” and “CHR-P syndrome” are used to refer to individuals who met criteria for a psychosis-risk syndrome on the Structured Interview for Psychosis-Risk Syndromes (SIPS).

### Referral

Family members, outpatient and inpatient clinical providers, pediatricians, school counselors or patients completed a standardized referral form. A licensed clinical psychologist or board-certified child and adolescent psychiatrist reviewed referral information and relevant psychiatric records. Individuals who: 1) were 12 to 30-years-old at time of referral, 2) were determined to have symptoms concerning for attenuated psychosis, 3) had program-accepted insurance, and 4) could attend an in-person/virtual evaluation in Massachusetts, were offered a diagnostic evaluation. Individuals who were determined to: 1) require other specialized services or a higher level of care at referral, 2) have a known diagnosis of a primary psychotic disorder (except Attenuated Psychosis Syndrome) or a history of experiencing full-threshold psychotic symptoms, 3) have an unstable or untreated severe eating disorder diagnosis, 4) have an intellectual disability (IQ≤70), and/or 5) a severe autism spectrum disorder (ASD) diagnosis, were provided alternative referrals.

### Diagnostic Evaluation and SIPS

Eligible individuals and their caregivers completed an online clinical assessment battery of self-report measures (see measures description below) and met with a child, adolescent, and adult psychiatrist (J.A.C.) for a 90-minute diagnostic evaluation. Collateral information from current treatment providers and prior medical records were reviewed as part of the evaluation. A SIPS assessment was recommended for patients for whom there were concerns, revealed by the self-report measures and/or diagnostic evaluation, for the presence of attenuated psychosis. The SIPS assessment was completed by a clinical psychologist or supervised psychology doctoral trainee who had been trained and certified on the SIPS. Patients who met SIPS criteria for Brief Intermittent Psychosis Syndrome (BIPS), Attenuated Positive Symptom Syndrome (APSS), or Genetic Risk and Functional Decline Syndrome (GRDS) with status of current progression, current persistence, or partial remission were considered to meet criteria for CHR-P syndrome. A feedback session for patients and their caregivers was co-led by the psychiatrist who conducted the initial evaluation and the psychologist or psychology trainee who administered the SIPS. During the feedback session, diagnostic impressions were shared. Recommendations were provided for further evaluation (e.g., additional medical evaluation, a magnetic resonance imaging brain scan, neuropsychological testing) and/or additional treatment (e.g., individual/group therapy, substance use treatment, school-based resources, psychopharmacology, and/or supported employment/vocational rehabilitation services). Patients who met criteria for CHR-P, resided in Massachusetts full-time, had insurance accepted by the program, and for whom staff were available to meet treatment needs were offered ongoing care in RE-SET. Translation services were used for the evaluation and feedback session as needed. Written reports from the diagnostic consultation were provided to patients, and their caregivers and current treatment providers with appropriate permission from the patient.

### Measures

We used the following reliable, validated, English-language instruments to assess psychotic symptoms, functioning, and clinical history. Attention was paid to selecting measures that were validated and reliable for ages 12+ years. Study data were collected online and managed using REDCap electronic data capture tools hosted at Massachusetts General Brigham (Harris et al., 2019).

The person making the referral reported on the patient’s lifetime history of violence (e.g. suicide, self-harm, aggression), substance use, psychiatric diagnosis, and psychiatric service use history. The referrer also completed the Adolescent Psychotic-like Symptom Screener (APSS; Kelleher et al., 2011), a 7-item scale assessing for current and past year psychotic experiences (yes/no). Endorsing more than two psychotic experiences on the APSS meets screening threshold for a possible psychosis spectrum illness (Kelleher et al., 2011). As part of the diagnostic evaluation, patients completed self-report measures on positive and negative psychotic symptoms; mood and anxiety symptoms; stress and traumatic experiences; behavioral difficulties; and social cognition. See Supplemental Table 1.

### Statistical Analysis

Descriptive statistics were used to characterize the sample. Univariate statistical tests (chi-square test or independent samples t-test) were used to test for differences in demographic and clinical variables at screening and diagnostic evaluation between patients who met CHR-P criteria and those that did not meet CHR-P criteria nor had concerns for a primary psychotic disorder (“non-CHR-P”). Statistical significance was two-tailed at an alpha level of .05. Effect sizes (Cramer’s V for chi-square test and Cohen’s d for t-test) were calculated, with magnitudes interpreted according to Cohen’s benchmarks (small: ≤ 0.5; medium: 0.5 - 0.8; large: ≥0.8) (Cohen, 1988).

## Results

Of the 118 help-seeking adolescents and young adults referred from October 2021 to December 2023, a total of 99 (mean age: 18 years; SD: 4) had complete referral data and were included in the analysis (see Table 1). Of the 57/99 (57.6%) screened to be eligible for diagnostic evaluation, 49/57 (86.0%) received a diagnostic evaluation, 48/49 (98.0%) completed the clinical assessment battery, and 31/49 (65.3%) completed the SIPS. Of those completing a diagnostic evaluation, 17/49 (34.7%) were not offered the SIPS due to presenting clinical concerns being better accounted for by non-psychotic disorders, or because the diagnostic evaluation was consistent with a full threshold psychotic disorder. Of those who received a SIPS interview, 24/31 (77.4%) met CHR-P criteria. See Figure 1 for patient eligibility flow diagram.

**Figure 1.**
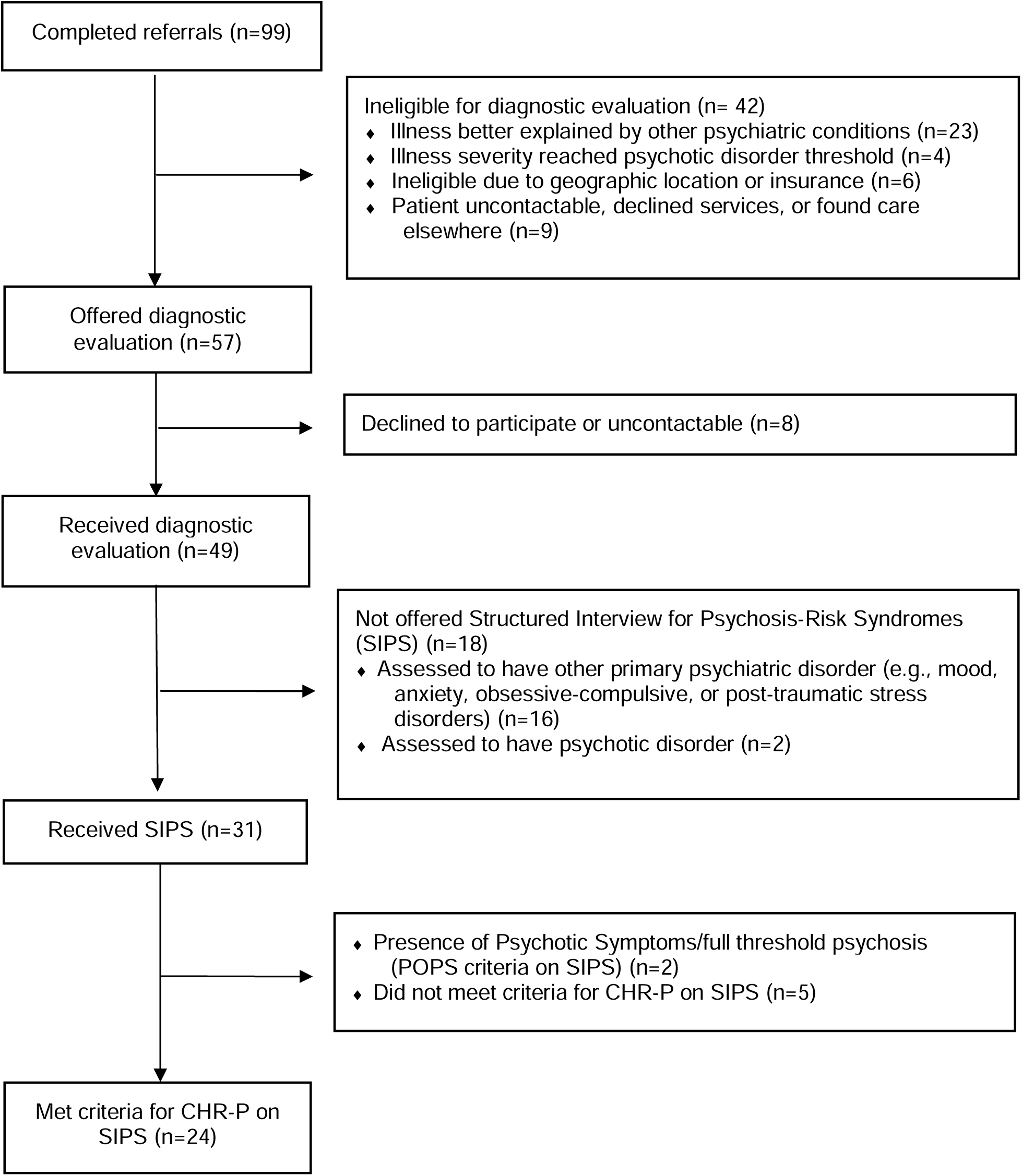
Flow Diagram of RE-SET Patient Eligibility for Assessment and Enrollment.

**Table 1.** Demographic and clinical characteristics at referral associated with Clinical High-Risk for Psychosis (CHR-P) (N=99).

| Referral Variable | Full Referral Sample (N = 99) | CHR-P (N = 24) | Non-CHR-P<sup>a</sup> (N = 75) | Statistic | p-value |
| --- | --- | --- | --- | --- | --- |
| Mean Age ± SD (Years) | 17.8 ± 4.0 | 17.4 ± 2.8 | 17.9 ± 4.0 | t(61) = -0.7 | .48 |
|  | N (%) | N (%) |  |  |  |
| Completed referral form | | | | $\chi^2(3)=0.19$ | .98 |
| Self | 17 (17.3) | 4 (16.7) | 13 (17.6) |  |  |
| Family member | 42 (42.9) | 11 (45.8) | 31 (41.9) |  |  |
| Outpatient provider<sup>b</sup> | 29 (29.6) | 7 (29.2) | 22 (30.9) |  |  |
| Acute psychiatric setting<sup>c</sup> | 10 (10.2) | 2 (8.3) | 8 (11.1) |  |  |
| Male Sex Assigned at Birth | 44 (44.3) | 12 (50) | 35 (42.7) | $\chi^2(1)=0$ | .96 |
| Gender Identity | | | | $\chi^2(6)=3.7$ | .71 |
| Male | 43 (43.4) | 10 (41.7) | 33 (45.1) |  |  |
| Female | 41 (41.4) | 9 (37.5) | 32 (41.5) |  |  |
| Non-binary | 7 (7.1) | 3 (12.5) | 4 (6.1) |  |  |
| Transgender male | 5 (5.1) | 1 (4.2) | 4 (4.9) |  |  |
| Transgender female | 2 (2.0) | 1 (4.2) | 1 (1.2) |  |  |
| Race/Ethnicity | | | | $\chi^2(6)=8.6$ | .20 |
| Asian/Pacific Islander | 8 (7.5) | 0 (0.0) | 8 (11.0) |  |  |
| Black/African American | 7 (6.6) | 2 (8.3) | 5 (7.3) |  |  |
| Hispanic or Latinx | 16 (15.1) | 1 (4.2) | 15 (19.5) |  |  |
| Middle Eastern or Northern African | 1 (0.9) | 0 (0.0) | 1 (2.4) |  |  |
| White | 56 (52.8) | 17 (70.8) | 39 (51.2) |  |  |
| Multiracial or Other | 10 (9.4) | 4 (16.7) | 6 (7.3) |  |  |
| English as Primary Language | 96 (97.0) | 24 (100) | 72 (67.9) | $\chi^2(4)=0$ | 1.0 |
| Lifetime Behaviors |  |  |  |  |  |
| Substance use | 35 (35.4) | 7 (29.2) | 28 (37.3) | $\chi^2(1)=0.23$ | .63 |
| Cannabis use | 33 (33.3) | 7 (29.2) | 26 (34.7) | $\chi^2(1)=0.06$ | .80 |
| Self-harm | 47 (47.5) | 12 (50) | 35 (46.7) | $\chi^2(1)=0.00$ | .96 |
| Suicide attempt | 28 (28.3) | 6 (25) | 22 (29.3) | $\chi^2(1)=0.02$ | .88 |
| Violence/aggression | 14 (14.1) | 3 (12.5) | 11 (14.7) | $\chi^2(1)=0$ | 1.0 |
| Criminal justice involvement | 4 (4.8) | 0 (0.0) | 4 (6.2) | NA |  |
| Lifetime Psychiatric Hospitalization | 51 (48.6) | 9 (37.5) | 42 (51.9) | $\chi^2(1)=1.8$ | .18 |
| Lifetime Psychiatric Diagnosis |  |  |  |  |  |
| Any | 82 (82.8) | 20 (80) | 63 (84) | $\chi^2(1)=0.0$ | 1.0 |
| Psychosis-spectrum disorders | 7 (7.1) | 3 (12.5) | 4 (5.3) | $\chi^2(1)=0.54$ | .46 |

ADVANCING ASSESSMENT OF CLINICAL HIGH-RISK FOR PSYCHOSIS
| Referral Variable | Full Referral Sample (N = 99) | CHR-P (N = 24) | Non-CHR-P<sup>a</sup> (N = 75) | Statistic | p-value |
| --- | --- | --- | --- | --- | --- |
| Mood disorders | 61 (61.6) | 14 (58.3) | 47 (62.7) | $\chi^2(1)=0.02$ | .89 |
| Anxiety disorders | 51 (51.5) | 14 (58.3) | 37 (49.3) | $\chi^2(1)=0.28$ | .59 |
| Obsessive-compulsive disorders | 15 (15.2) | 3 (12.5) | 12 (16) | $\chi^2(1)=0.01$ | .93 |
| Posttraumatic stress disorders | 11 (11.1) | 3 (12.5) | 8 (10.7) | $\chi^2(1)=0.0$ | 1.0 |
| Eating disorders | 4 (4.0) | 1 (4.2) | 3 (4.0) | $\chi^2(1)=0.0$ | 1.0 |
| Personality disorders | 4 (4.0) | 1 (4.2) | 3 (4.0) | $\chi^2(1)=0.0$ | 1.0 |
| Attention-deficit/hyperactivity disorder | 31 (31.3) | 10 (41.7) | 21 (28.0) | $\chi^2(1)=1.1$ | .32 |
| Autism Spectrum Disorders | 11 (11.1) | 6 (25) | 5 (6.7) | $\chi^2(1)=4.5$ | **.03** |
| Have Current Outpatient Mental Health Provider |  |  |  |  |  |
| Any | 79 (79.8) | 19 (79.2) | 60 (80) | $\chi^2(1)=0$ | 1.0 |
| Prescriber<sup>d</sup> | 61 (61.6) | 13 (54.2) | 48 (64.0) | $\chi^2(1)=0.39$ | .53 |
| Therapist | 53 (50.5) | 14 (58.3) | 39 (48.1) | $\chi^2(1)=0.1$ | .75 |
| School counselor | 29 (29.3) | 7 (29.2) | 22 (27.2) | $\chi^2(1)=0$ | 1.0 |
| Current IEP, 504 Plan, or College Accommodations | 40 (40.4) | 12 (50) | 28 (37.0) | $\chi^2(1)=0.74$ | .39 |
| Current Psychotropic Medication | 73 (73.7) | 18 (75) | 52 (69.3) | $\chi^2(1)=0.4$ | .51 |
| Current Antipsychotic Medication | 38 (38.3) | 5 (20.8) | 31 (41.3) | $\chi^2(1)=6.8$ | **.01** |
| Family History of Psychiatric Illness |  |  |  |  |  |
| Any | 68 (68.7) | 16 (66.7) | 52 (69.3) | $\chi^2(1)=0$ | 1.0 |
| Psychotic disorders | 31 (31.3) | 6 (25) | 25 (33.3) | $\chi^2(1)=0.26$ | 0.61 |
| Adolescent Psychotic-like Symptom Screener (APSS) |  |  |  |  |  |
| APSS Mean Total Score (SD) | 1.9 ± 1.5 | 2.2 ± 1.2 | 1.7 ± 1.5 | t(41.3) = 1.4 | .18 |
| Met APSS >1 cut-off | 47 (57.3) | 16 (80.0) | 31 (50) | $\chi^2(1)=4.4$ | **.04** |

<sup>a</sup> No concern for primary psychotic disorder or CHR-P, or did not meet criteria for CHR-P on the SIPS.
<sup>b</sup> Outpatient provider included primary care physicians, pediatricians, outpatient mental health providers, school counselors/educators.
<sup>c</sup> Acute psychiatric setting included emergency departments and inpatient/partial psychiatric hospitals.
<sup>d</sup> Psychiatric prescriber, pediatrician, or primary care provider prescribing psychotropic medication.

### Patient characteristics at referral and associations with CHR-P syndrome (Table 1)

Referred patients (N= 99) had high rates of psychiatric comorbidity at the time of referral. A large proportion (82.8%) reported at least one psychiatric diagnosis; more than half reported having been diagnosed with a mood (61.6%) or anxiety disorder (51.5%). Close to 80% of patients were typically in current mental health treatment; 68.9% reported current psychotropic medication treatment, including over one-third (35.8%) reporting current antipsychotic medication treatment. Referred patients also had high psychiatric acuity and risk; of these patients, 48.6% had a previous psychiatric hospitalization, 28.3% had a past suicide attempt, 47.5% had engaged in self-harm-related behaviors, and 35.4% had used substances (tobacco, alcohol, cannabis). More than half (57.3%) of the referred patients met the screening cut-off for likely psychosis spectrum illness on the APSS.

Compared to those who did not meet criteria for CHR-P syndrome, significantly more individuals who met CHR-P criteria had a referrer-reported history of ASD diagnosis (CHR-P, N=24: 25% vs. non-CHR-P, N= 75: 6.7%; χ^2^(1) = 4.5, Cramér’s V = 0.21, p =.03) and met the APSS screening cut-off (CHR-P: 80% vs. non-CHR-P: 50%; χ^2^(1) = 4.4, Cramér’s V = 0.23, p =.04). CHR-P individuals were also less likely to report a history of antipsychotic medication treatment (CHR-P: 20.8% vs. non-CHR-P: 41.3%; χ^2^(1) = 6.8, Cramér’s V = 0.33, p = .01). These factors had small effect sizes on predicting group differences.

### Antipsychotic medication in non-CHR sample

Given our finding that referrer-reported current antipsychotic medication was higher in the group of patients that did not meet CHR-P criteria compared to the group that did, we conducted *post hoc* analyses to test for differences within the non-CHR-P group between those who were (N=44) and were not currently treated with antipsychotic medications (N=31). Compared to non-CHR-P patients who had no history of antipsychotic medication treatment, non-CHR-P patients who were treated with antipsychotic medications were more likely to have a history of violent behaviors (29.0% vs. 4.5%, χ^2^ (1) = 6.9, Cramér’s V = 0.30, p =.009) and a history of psychiatric hospitalizations (80.6% vs. 38.6%, χ^2^ (1) = 11.4, Cramér’s V = 0.39, p <.001). There were no significant associations between antipsychotic treatment in the non-CHR-P group at time of referral and other demographic factors, substance use history, self-harm and suicidal behavior history, criminal justice involvement history, or family history of psychiatric or psychotic illness (all p > 0.2).

### Clinical characteristics at diagnostic evaluation and associations with CHR-P syndrome (Table 2)

Patients who received a diagnostic evaluation (N = 49) had high scores on measures assessing positive psychotic symptoms (Brief State Paranoia Checklist and Prodromal Questionnaire - Brief) (See Supplemental Table 1 for scoring interpretation). Consistent with the transdiagnostic nature of the CHR-P population, patients meeting CHR-P criteria reported high levels of negative symptoms, including deficits in anticipatory pleasure and social withdrawal (preferring substantial time alone), as well as moderate levels of loneliness, perceived stress, and depression; moderate exposure to emotional abuse and neglect; mild to moderate deficits in social interaction; and severe anxiety symptoms. However, there were no significant differences between those who met CHR-P syndrome and those who did not on these measures (all p > 0.10).

**Table 2.** Descriptives and statistical tests for clinical characteristics associated with Clinical High-Risk for Psychosis (CHR-P) at diagnostic evaluation (N = 48).

| Measure | M (SD) / N (%) |  | Statistic | d [95% CI] | P-value |
| --- | --- | --- | --- | --- | --- |
|  | CHR-P<br>(N=21) | Non-CHR-P<sup>a</sup><br>(N=27) |  |  |  |
| **Self-Report** |  |  |  |  |  |
| Prodromal Questionnaire - Brief | 11.9 (5.9) | 8.9 (6.3) | t(43.5) = 1.64 | 0.48<br>[-0.11, 1.07] | .11 |
| Paranoid Checklist 5-item Scale | 13.4 (9.1) | 12.6 (9.1) | t(43.2) = 0.28 | 0.08<br>[-0.49, 0.65] | .78 |
| Temporal Experience of Pleasure Scale |  |  |  |  |  |
| Anticipatory pleasure | 38.4 (10.4) | 36.3 (10.8) | t(44) = 0.68 | 0.20<br>[-0.37, 0.77] | .50 |
| Consummatory pleasure | 31.6 (8.0) | 30.3 (8.5) | t(44) = 0.53 | 0.16<br>[-0.42, 0.74] | .60 |
| UCLA Loneliness Scale | 47.3 (14.0) | 45.7 (11.6) | t(38.7) = 0.44 | 0.13<br>[-0.44, 0.71] | .66 |
| Time Alone Questionnaire |  |  |  |  |  |
| Hours awake per day | 17.6 (1.2) | 16.0 (1.5) | t(40.9) = 3.73 | **1.12**<br>**[0.46, 1.78]** | **< .001** |
| Time spent alone (% of hours awake) | 60.8 (27.1) | 53.6 (29.7) | t(38.7) = 0.83 | 0.25<br>[-0.36, 0.86] | .41 |
| Time preferred alone (% of hours awake) | 57.2 (28.5) | 62.3 (26.5) | t(35.2) = -0.59 | -0.18<br>[-0.80, 0.43] | .56 |
| Beck Depression Inventory-II | 20.8 (13.3) | 23.3 (14.8) | t(43.8) = -0.60 | -0.18<br>[-0.76, 1.07] | .55 |
| Emotion Reactivity Scale | 39.4 (24.6) | 44.7 (23.4) | t(42.0) = -0.76 | -0.22<br>[-0.80, 0.36] | .45 |
| Perceived Stress Scale | 22.2 (7.2) | 23.0 (7.6) | t(40.1) = -0.35 | -0.11<br>[-0.70, 0.49] | .73 |
| Childhood Trauma Questionnaire |  |  |  |  |  |
| Emotional Abuse | 9.5 (5.0) | 11.9 (5.2) | t(43.3) = -1.56 | -0.46<br>[-1.05, 0.13] | .13 |
| Physical Abuse | 5.8 (1.2) | 6.8 (2.7) | t(34.2) = -1.62 | -0.45<br>[-1.04, 0.14] | .12 |
| Emotional Neglect | 11.1 (5.3) | 11.6 (4.7) | t(40.5) = -0.28 | -0.08<br>[-0.66, 0.50] | .78 |
| Physical Neglect | 10.1 (2.0) | 10.4 (2.1) | t(43.2) = -0.65 | -0.19<br>[-0.77, 0.39] | .52 |
| Minimization/Denial | 8.3 (4.3) | 8.0 (3.9) | t(41.1) = | 0.08 | .79 |

ADVANCING ASSESSMENT OF CLINICAL HIGH-RISK FOR PSYCHOSIS
| Measure | M (SD) / N (%) |  | Statistic | *d* [95% CI] | *p*-value |
| --- | --- | --- | --- | --- | --- |
|  | CHR-P<br>(N=21) | Non-CHR-P<sup>a</sup><br>(N=27) |  |  |  |
|  |  |  | 0.27 | [-0.50, 0.66] |  |
| Conners-Wells Adolescent Self-Report Scale (Short Form) |  |  |  |  |  |
| Cognitive Problems | 9.8 (4.9) | 6.6 (4.4) | t(36.6) = 2.28 | **0.70**<br>**[0.08, 1.31]** | **.03** |
| Hyperactivity | 10.2 (4.1) | 7.0 (4.1) | t(38.5) = 2.56 | **0.78**<br>**[0.16, 1.40]** | **.02** |
| Conduct Problems | 3.5 (1.8) | 2.8 (2.5) | t(41.7) = 0.99 | 0.29<br>[-0.31, 0.88] | .33 |
| ADHD | 11.0 (5.3) | 9.9 (5.6) | t(39.9) = 0.67 | 0.20<br>[-0.40, 0.80] | .51 |
| Mentalization Scale - Total | 90.9 (11.9) | 93.7 (13.9) | t(41.3) = -0.72 | -0.21<br>[-0.81, 0.39] | .48 |
| Others | 32.4 (6.1) | 35.8 (5.4) | t(36.5) = -1.89 | -0.58<br>[-1.19, 0.03] | .07 |
| Self | 22.3 (6.0) | 23.9 (7.9) | t(42.0) = -0.79 | -0.23<br>[-0.83, 0.37] | .43 |
| Motivation | 36.2 (5.6) | 34.0 (6.5) | t(41.3) = 1.21 | 0.36<br>[-0.24, 0.96] | .23 |

<sup>a</sup> No concern for primary psychotic disorder or CHR-P, or did not meet criteria for CHR-P on the SIPS.

CHR-P individuals reported spending significantly more hours awake (17.6 ± 1.2 hours) than those who did not meet CHR-P criteria (16.0 ± 1.5 hours), a difference that was associated with a large effect size (d [95% CI] = 1.12 [0.46, 1.78], p < 0.001). They also self-reported significantly more cognitive difficulties (Conners-Wells Adolescent Self-Report Scale – Short Form, cognitive problems score: 9.8 ± 4.9) than those who did not meet CHR-P criteria (6.6 ± 4.4), with a medium effect size (d = 0.70 [0.08, 1.31], p = 0.03). Additionally, CHR-P individuals reported significantly more behavioral hyperactivity (Conners-Wells Adolescent Self-Report Scale – Short Form, hyperactivity score: 10.2 ± 4.1) than those who did not meet CHR-P criteria (7.0 ± 4.1), with a medium effect size (d = 0.78 [0.16, 1.40], p = 0.02).

## Discussion

This study provides a description and initial empirical evidence supporting the clinical utility of specialized CHR-P evaluation and treatment programs built upon learning health system principles. The RE-SET Program demonstrates the feasibility of integrating standardized data collection into routine clinical practice, presenting a model for addressing the complex needs of at-risk individuals with attenuated psychotic symptoms, while simultaneously advancing scientific understanding of psychosis risk and bridging the research-to-practice gap. Using real-world data, our study highlights that individuals who are evaluated for CHR-P present with clinical and diagnostic complexity and multidimensional risk factors. Notably, individuals who met criteria for CHR-P on the SIPS were more likely to report a history of an ASD diagnosis, reduced sleep duration, more cognitive difficulties and more hyperactivity relative to individuals who did not meet CHR-P criteria. The identification of these specific predictors associated with CHR-P syndrome provides data-driven guidance for clinical decision-making, enabling research findings to be practically applied to improve the screening, evaluation, and treatment of CHR-P.

Notably, we identified neurodevelopmental, symptomatic, and behavioral indicators that distinguished individuals who met criteria for CHR-P syndrome among a help-seeking sample. Individuals who met CHR-P criteria had more self-reported cognitive difficulties and attention symptoms than those who did not meet CHR-P criteria. Cognitive impairments in CHR-P are associated with poor functioning and an increased risk for transition to psychosis (Catalan, Salazar De Pablo, et al., 2021; Seidman et al., 2016). Interestingly, our findings are consistent with the growing evidence for the overlap and high comorbidity between autism and psychosis spectrum disorders (Lugo Marín et al., 2018; Solomon et al., 2011). Also, emerging research has increasingly highlighted sleep disruption as a potential prodromal marker of psychotic illnesses, which has the potential to precipitate and worsen psychotic symptoms (Reeve et al., 2019).

These factors together reflect the complex developmental, cognitive, and behavioral profile associated with attenuated psychotic symptoms and psychosis-risk. These findings suggest that comprehensive screening and evaluation for CHR-P can be improved by incorporating sensitive assessment of psychotic symptoms, developmental history, evaluation of sleep patterns, and cognitive and behavioral functioning assessment.

The transdiagnostic clinical characteristics and risk factors associated with CHR-P syndrome align with the emerging perspective that CHR-P syndrome is not solely a precursor to primary psychotic disorders but represents a broader syndrome of psychological vulnerability to a wide range of serious mental illnesses and functional impairment (McGorry et al., 2018). Subclinical psychotic symptoms, in particular, are not only associated with an elevated risk for developing a psychotic disorder (Hodgekins et al., 2018), but also increase risk of developing anxiety, mood, and substance use disorders, and a more severe illness course (Hodgekins et al., 2018; van Os, 2013; Wigman et al., 2012). Identification of the CHR-P syndrome should ideally lead to referral to not only specialized CHR-P treatment, but also other appropriate youth mental health services to prevent clinical deterioration and poor outcomes in help-seeking youth presenting with high clinical need.

Individuals meeting CHR-P criteria were less likely to be treated with antipsychotic medications, compared to those who did not meet CHR-P criteria. Individuals prescribed antipsychotics without CHR-P had more severe psychiatric illness, as evidenced by more violent behaviors and psychiatric hospitalizations. However, it is also possible that treatment with antipsychotics (for a variety of indications) masked the presence of underlying psychosis. In children and adolescents, antipsychotics are often used off-label to treat a wide range of behavioral disturbances and are not selectively used to treat psychotic disorders (CMS, 2015).

These findings, combined with prior observational studies showing greater antipsychotic treatment in CHR-P individuals who transitioned to psychosis (Raballo et al., 2020, 2021; Zhang et al., 2020), suggest that antipsychotic treatment may be a general marker of psychiatric illness severity. Greater availability of specialized and intensive treatment settings is likely necessary to reduce antipsychotic prescriptions and provide alternatives to antipsychotic treatment of severe behavioral dysregulation.

Further, these CHR-P predictors and co-occurring clinical characteristics across developmental, symptomatic, and functional domains underscore the need for transdiagnostic approaches to CHR-P intervention as well. Such a population would benefit from a stepped model of care, where those who were initially found not to meet CHR-P criteria continue to be monitored and assessed for CHR-P syndrome and other psychiatric illnesses (e.g., annually) and access general youth mental health services (McGorry et al., 2022). Those who do meet CHR-P criteria can receive intensive and specialized coordinated specialty care for CHR-P, such as that provided by the RE-SET Program. Specialized CHR-P treatment programs will require competent clinicians who are skilled in the transdiagnostic assessment and treatment of psychosis and distinct and co-occuring symptoms groups including ASD. Evidenced-based treatments would need to address difficulties with emotion regulation, self-harm, suicidality, substance use, sleep, social functioning and cognition. Consistent with the therapeutic model employed in RE-SET, CHR-P treatment can implement an integrated toolkit of evidence-based interventions targeted at transdiagnostic risk factors such as resilience, emotional avoidance (Barlow et al., 2017), emotion regulation, social cognition (Burke et al., 2020; Clauss et al., 2025; DeTore et al., 2023), and substance use (McCarthy et al., 2022). Thoughtful prescribing and de-prescribing of anti-psychotics is also a critical component of CHR-P treatment.

### Limitations

This study has several limitations. Patients were selected based on referrals to the RE-SET Program, which may limit the generalizability of the findings to other CHR-P samples. Also, several measures were only completed by a subset of the patients, as the clinical evaluation process was revised and streamlined during the study period, resulting in measures being added or removed. Patients who had difficulty completing online self-report questionnaires prior to evaluation were offered the opportunity to complete questionnaires in-person at the RE-SET Program; however, not all patients completed the full battery of questionnaires. The data reported were reliant on referrer-report, self-report, and clinical interviews. Additionally, cognitive measures were based on self-report of cognitive functioning as opposed to neuropsychological testing. Longitudinal follow-up assessments were not available at the time of data analysis but are currently ongoing.

### Future Directions

Our findings, from one CHR-P program, have informed the development and refinement of a CHR-P evaluation process and battery based on multidimensional risk factors. This battery has served as the model for standardizing a core assessment battery across all CHR-P clinics that are part of a larger, early psychosis-focused learning health system in Massachusetts. With ongoing data collection integrated with clinical care, future investigations will be focused on longitudinal tracking of clinical characteristics to understand illness and recovery trajectories, and clinical outcomes following CHR-P treatment, informing the development of treatment algorithms based on different risk profiles. By employing longitudinal measurement-based care, we can also begin to develop personalized and effective treatment plans that optimize outcomes. Technological advances, such as wearables for measuring sleep and activity levels (Fonseka & Woo, 2022), computer-based neuropsychological assessments (Singh et al., 2021), and virtual reality-based interventions (Holt et al., 2025), may contribute to risk assessment and treatment algorithms in the future.

## Conclusions

Individuals referred for CHR-P evaluation present with diagnostic complexity and significant clinical need. This complex clinical profile requires a comprehensive, transdiagnostic approach to screening, assessment, and intervention, with attention to a broad range of psychiatric symptoms, cognitive, behavioral, and sleep difficulties, and co-occurring ASD. A specialized CHR-P evaluation and treatment program that incorporates routine clinical assessment is feasible and valuable, providing data-driven opportunities for improving identification of CHR-P syndrome, selecting treatment targets, and personalizing treatment to improve outcomes for individuals with CHR-P.

### Key points and relevance

**What’s known:**

- Clinical high-risk for psychosis (CHR-P) represents a critical opportunity for prevention of serious mental illnesses, but care models for evaluating and treating a complex and heterogenous population are limited.

**What’s new**:

- Learning health system principles can be applied to improve assessment and treatment of CHR-P as part of a specialized clinical service.
- History of autism spectrum disorder, more attenuated psychotic symptoms, reduced sleep duration, and cognitive and behavioral difficulties may be more common in those who meet criteria for CHR-P than in those who do not.

**What’s relevant**:

- CHR-P services should comprehensively assess developmental history, sleep patterns, and cognitive and behavioral functioning, alongside gold-standard psychosis risk assessment.
- Routine measurement-based care enables continuous improvement in assessment and intervention protocols, providing opportunities for not only personalized treatment, but also for quality improvement efforts for service delivery models.
- Current treatment with anti-psychotic medication may indicate more severe psychiatric illness and should not necessarily be an exclusion for CHR-P care.

## Data Availability

All data produced in the present study are available upon reasonable request to the authors.

## Acknowledgements

**Study funding:** Dr. Foo was supported by funding from the Massachusetts Department of Mental Health to the Massachusetts General Hospital Center of Excellence for Psychosocial and Systemic Research, and the Sidney R. Baer, Jr Foundation. Dr. Donovan was supported by the Massachusetts Department of Mental Health. Dr. Cather was supported by funding from the Massachusetts Department of Mental Health to the Massachusetts General Hospital Center of Excellence for Psychosocial and Systemic Research. Dr. Holt was supported by funding from the National Institute of Mental Health (R01MG127265; R01MH125426; and R01MH122371-SCH), the Massachusetts Department of Mental Health, and the Sidney R. Baer, Jr Foundation. Dr. Clauss was supported by the Dupont Warren Fellowship of Harvard Psychiatry, Louis V. Gerstner Scholar Award, the Chen Institute Mass General Neuroscience Transformative Scholar Award, and the Maryland Psychiatric Research Center. **Contributorships:** All authors contributed to the design of the study and participated in review of the data and manuscript. Dr. Clauss and Dr. Foo had full access to the data in the study and take responsibility for the integrity of the data in the study and the accuracy of the data analysis. The authors thank Drew Coman, PhD, Alexandra Volpacchio, PsyD, Victoria Hasler, PhD, and psychology doctoral trainees for conducting SIPS assessments; Tvisha Chatterjea for data curation; and Kevin Potter, PhD, for consultation on statistical analysis. The authors thank the patients and families who were evaluated and participated in data collection. **Conflicts of interest**: The authors report no conflicts of interest. Dr. Donovan’s spouse is an investor in Mirah, a privately-held measurement-based health care company.

## Abbreviations

*CHR-P*: clinical high-risk for psychosis
*RE-SET*: Resilience Evaluation-Social Emotional Training
*SIPS*: Structured Interview for Psychosis-Risk Syndromes
*ASD*: autism spectrum disorder

