## SupplementalMaterial for "Implementing Learning Health System Principles to Advance the Evaluation and Treatment of Clinical High-Risk for Psychosis"

**Supplemental Table 1. *Patient self-report measures used in clinical assessment battery.***

| Construct | Self-Report Measure | Measure Description | Number of items (score range) | Scoring interpretation |
| --- | --- | --- | --- | --- |
| Positive psychotic symptoms | Brief State Paranoia Checklist (PC-5) (Schlier et al., 2016) | PC-5 is a shortened version of the Paranoia Checklist (PCL) designed to assess state-level paranoid ideation that fluctuates over time. Participants rate level of agreement with statements related to paranoid beliefs on a 7-point Likert scale. | 5 (0-35) | Higher total scores indicate higher levels of paranoia. |
|  | Prodromal Questionnaire – Brief (PQ-B) (Loewy et al., 2011) | PQ-B was developed from the original 92-item Prodromal Questionnaire to screen for psychosis risk syndromes. Participants rate whether they have experienced various positive psychotic symptoms in the past month (Yes/No). If they endorse symptoms, participants rate extent to which symptoms cause distress or functional impairment on a 5-point Likert scale. | 21 (0-21 symptoms endorsed; 21 – 105 level of distress/impairment) | Total of ≥3 positive symptoms or ≥6 total distress score sensitively identifies likely CHR-P individuals. |
| Negative psychotic symptoms | Temporal Experience of Pleasure Scale (TEPS) (Gard et al., 2006) | TEPS is developed to measure individual trait dispositions in both anticipatory and consummatory experiences of pleasure. Participants rate extent to which they agree with statements on a 6-point Likert scale. | 10 (anticipatory pleasure scale; 10-60); 8 (consummatory pleasure scale; 8-48) | Lower total scores indicate more deficits. |
|  | UCLA Loneliness Scale (Russell, 1996) | The scale is designed to measure one’s subjective feelings of loneliness and feelings of social isolation. Participants rate how often they feel lonely or social isolated on a 4-point Likert scale. | 20 (20 – 80) | Total scores of 35-49 indicate moderate degree of loneliness, ≥50 indicate high degree of loneliness. |
|  | Time Alone Questionnaire (TAQ) (Zapetis et al., 2022) | This scale evaluates how much of the day individuals spend asleep, how much alone and with others, and the percentage of the day they’d prefer to spend alone versus with others. | 4 (0 – 24 hours; 0-100%) | Larger % of the day preferred alone indicates higher degree of social anhedonia |
| Mood and anxiety symptoms | Beck Depression Inventory-II (BDI-II) (Beck et al., 2011) | This scale measures the severity of depression in adolescents and adults, consistent with the 4^th^ Diagnostic and Statistical Manual of Mental Disorders (DSM-IV) criteria for major depressive disorder. For various depressive symptoms, participants pick out the statement that best describes the way they have felt over the past two weeks. Statements are ranked by severity of symptom on a 4-point Likert scale. | 21 (0 – 63) | Total scores of 20-28 indicate moderate depression symptoms, 29-63 indicate severe depression symptoms. |
|  | Emotion Reactivity Scale (ERS) (Nock et al., 2008) | The ERS is designed to assess individual’s experience of emotion reactivity on a regular basis. Participants rate extent to which statements about emotional sensitivity (e.g., “I tend to get emotional very easily.”), arousal/intensity (“When I experience emotions, I feel them very strongly/intensely”), and persistence (“When I am angry/upset, it takes me much longer than most people to calm down”) describe them on a 5-point Likert scale. | 21 (0 – 44) | Higher total scores indicate higher emotional reactivity |
| Stress and traumatic experiences | Perceived Stress Scale (PSS) (Cohen et al., 1983) | PSS is the most widely used instrument for measuring the degree to which situations in one’s life are appraised as stressful. Participants rate how often they have stressed from different situations in the past month on a 5-point Likert scale. | 10 (0 – 40) | Higher total scores indicate higher perceived stress. Total score of 0-13 would be considered low stress; 14-16 considered moderate stress; 27-40 considered high stress. |
|  | Childhood Trauma Questionnaire – Short Form (CTQ) (Bernstein et al., 1994) | This scale includes 25 clinical items measuring an adolescent’s or adult’s experiences of child abuse and neglect, retrospectively. The clinical items are separated into five subscales of five items each: emotional, physical, and sexual abuse, and emotional and physical neglect. The scale includes 3 validity items to detect the underreporting of maltreatment. Participants rate severity of abuse or neglect on a 5-point Likert scale. | 5 subscales of 5 item each (0-25) | Higher total scores indicate greater the severity of abuse, across five subscales. |
| Behavioral Problems | Conners-Wells’ Adolescent Self-Report Scale Short Form (CASS:Short) (Conners et al., 1997) | The CASS:Short is a 27-item self-report instrument derived from the CASS:Long (Conners et al., 1997) validated for 12-17 year-olds, which we used for all evaluated clients regardless of their age. The items make up three factors: conduct problems (likelihood of breaking rules and engaging in antisocial activities), cognitive problems (difficulty organizing and completing tasks), and hyperactivity (difficulty sitting still or doing same task for very long). It also includes an ADHD index to assess probable ADHD. Participants rate extent they agree with each statement on a 4-point Likert scale. | 27 (conduct problems, 6 items: 0-24; cognitive problems, 6 items: 0-24; hyperactivity, 6 items: 0-24; ADHD index, 12 items: 0-48) | Higher scores indicate more conduct, cognitive, hyperactivity problems, respectively. Higher scores on ADHD index indicate likely ADHD. |
| Social Cognition | Mentalization Scale (Dimitrijević et al., 2018) | This measure assesses mentalizing capacity in three dimensions: self-related mentalization, other-related mentalization, and motivation to mentalize. Participants rate degree to which they agree with statements on mentalization abilities on a 5-point Likert scale. | 10 (self; 10 – 50); 8 (other; 8 – 40); 10 (motivation; 10 – 50) | Higher total scores on each subscale indicate a more advanced capacity for mentalizing. |

**Supplemental Table 2. *Caregiver self-report measures used in clinical assessment battery and results.***

Caregivers reported on their loved one’s attentional and behavioral difficulties; autism spectrum disorder symptoms; and developmental history. No caregiver-reported clinical characteristics were significantly associated with CHR-P status.

| Construct | Caregiver- Report Measure | Measure Description | Number of items (score range) | Scoring interpretation |
| --- | --- | --- | --- | --- |
| Attentional and Behavioral Difficulties | Conners' Parent Rating Scale-Revised (CPRS-R) (Conners et al., 1998) | The CPRS-R is a caregiver report of common presenting problems for children (ages 3 to 17 years) referred to an outpatient psychiatric setting, focusing on behaviors directly related to attention deficit-hyperactivity disorder (ADHD). We included subscales of oppositional problems, cognitive problems, hyperactivity, and an ADHD index Raw scores were converted into T-scores based on norms for different age groups and sexes. | 57 | T-score ≤ 60 indicates no significant behavioral or academic problems. T-score ≥ 61-70 suggests  potential behavioral or academic issues. T-score ≥ 70 indicates more severe issues. Higher scores on the ADHD index likely ADHD. |
| Autism Spectrum Disorder Symptoms | Social Responsiveness Scale (SRS-P) (Constantino & Gruber, 2005) | The SRS is a 65-item parent-report for children and adolescents (ages 4-18) that measure the severity of Autism Spectrum symptoms as they occur in natural social settings. The SRS provides a clear picture of a child’s social impairments, assessing social awareness, social information processing, capacity for reciprocal social communication, social anxiety/avoidance, and autistic preoccupations and traits. Each item on the scale inquires about an observed aspect of reciprocity behavior. Behaviors are rated on a 4-point Likert scale. The raw scores are converted  into T-scores, based on norms for different age groups and sexes. | 65 | T-score ≤59 indicates behaviors within normal limits, generally not associated with ASD. T-score 60-65 indicates mild degree of social impairment. T-score 66-75 indicates moderate degree of social impairment. T-score ≥ 76 indicates severe degree of social impairment, most strongly associated with clinical diagnosis of ASD. |
| Developmental history | Caregiver report on clinical assessment battery | Caregiver also reported on history of pregnancy complications or prenatal exposure to substances, as well as age when patients met early motor milestones (month demonstrating ability to roll from tummy to back, lean on hands to support when sitting, sit without support by themselves; pull up to stand, and walk) and language and communication milestones (month demonstrating first word, two-word speech, three-word speech). |  | Normative data for individual milestones following the American Academy of Pediatrics and Centers for Disease Control and Prevention’s recommendations (Zubler et al., 2022) were used to determine if eligible patients exhibited any motor or language delays. |

| **Measure** | **M (SD) / N (%)** | | | **Statistic** | **d [95% CI]** | ***p-*value** |
| --- | --- | --- | --- | --- | --- | --- |
|  | **CHR-P (N=21)** | **Non-CHR-P (N=27)** | |  |  |  |
| ***Caregiver-Report*** |  |  |  | |  |  |
| Conners’ Parent Rating Scale (T-Score) |  |  |  | |  |  |
| Oppositional Problems | 56.8 (10.3) | 60.1 (15.6) | t(40.0) =  -0.85 | | -0.25 [-0.84, 0.35] | .40 |
| Cognitive Problems | 57.6 (10.3) | 61.5 (14.2) | t(42.6) =  -1.08 | | -0.31 [-0.90, 0.28] | .29 |
| Hyperactivity | 58.9 (17.4) | 60.3 (17.8) | t(41.2) =  -0.27 | | -0.08 [-0.67, 0.51] | .79 |
| ADHD | 7.9 (4.0) | 8.1 (4.8) | t(43.0) =  -0.18 | | -0.05 [-0.64, 0.54] | .86 |
| Social Responsiveness Scale – Total (T-Score) | 62.4 (9.0) | 65.0 (12.2) | t(42.8) =  -0.81 | | -0.23 [-0.82, 0.36] | .42 |
| Social awareness | 57.1 (9.2) | 58.8 (13.2) | t(43.6) =  -0.52 | | -0.15 [-0.73, 0.44] | .61 |
| Social cognition | 60.1 (11.5) | 60.0 (13.6) | t(42.0) = 0.04 | | 0.01 [-0.58, 0.61] | .97 |
| Social communication | 65.6 (6.8) | 66.4 (9.3) | t(43.9) =  -0.33 | | -0.09 [-0.68, 0.49] | .74 |
| Social motivation | 62.0 (10.3) | 67.9 (13.6) | t(44.0) =  -1.69 | | -0.49 [-1.07, 0.11] | .10 |
| Restricted and repetitive interest | 57.6 (11.5) | 64.2 (14.8) | t(43.0) =  -1.69 | | -0.49 [-1.09, 0.11] | .10 |
| Social communication and interaction | 65.0 (8.9) | 65.8 (11.3) | t(41.9) =  -0.28 | | -0.08 [-0.68, 0.51] | .78 |
| Any pregnancy complication | 1 (4.3) | 1 (1.3) | χ^2^ (1) = .12 | | - | .73 |
| Any prenatal exposure to substances | 2 (8.7) | 1 (1.3) | χ^2^ (1) = .95 | | - | .33 |
